# Whole exome-trio analysis reveals rare variants associated with Congenital Pouch Colon

**DOI:** 10.1101/2019.12.10.19013680

**Authors:** Sonal Gupta, Praveen Mathur, Ashwani Kumar Mishra, Krishna Mohan Medicherla, Prashanth Suravajhala

**Affiliations:** Department of Biotechnology and Bioinformatics, Birla Institute of Scientific Research (BISR), Statue Circle, Jaipur 302021, RJ India; Department of Biotechnology, Amity University Rajasthan, Kant Kalwar, Jaipur 303002, RJ India; Department of Pediatric surgery, SMS medical college and Hospital, JLN Marg, Jaipur 302004, RJ India; DNA Xperts, Noida, India

**Keywords:** whole exome sequencing, trio exome, missense, congenital pouch colon

## Abstract

Anorectal malformations (ARM) are individually common but Congenital Pouch Colon (CPC), a rare anorectal anomaly causes a dilated pouch in genitourinary tract. We have earlier attempted to understand the clinical genetic makeup of CPC and identified genes responsible for the disease using whole exome sequencing (WES). Here we report our studies of CPC, by identifying *de novo* heterozygous missense mutations in 16 proband-parent trios and further discover variants of unknown significance which could provide insights into CPC manifestation and its etiology. Our study confirms candidate mutations in genes, *viz*. C7orf57, C10orf120, C9orf84 and MUC16, CTC1 particularly emphasizing the role of hypothetical genes or open reading frames causing this developmental disorder. Variant validation revealed disease causing mutations associated with CPC and genitourinary diseases which could close the gaps of surgery in bringing intervention in therapies.

---

Whole Exome Sequencing (WES) has been an invaluable and cost-effective genomic approach to identify genetic variants responsible for both Mendelian and polygenic diseases (*Ng et al., 2010*). In the recent past, it has allowed detecting clinically relevant genomic regions spanning the known unknowns regions, disease-associated sites and untranslated regions (UTRs) (*Shen et al., 2014*). In addition to the well known diseases, prenatal abnormalities, structural anomalies and congenital defects were studied demonstrating a good diagnostic yield (*Greenbaum et al., 2019; Mone et al., 2018*). While WES approaches are abundant, they are limited if the disease in question is characteristically rare and medically inconclusive. This could be deterrent because of the challenges in variant discovery, including rare and low-frequency mutations using next generation sequencing (NGS) technologies. Recent decrease in cost of WES and accuracy of the NGS has enabled the researchers to study trios (proband/parents) or quad, in case of an addition of sibling to discover single nucleotide variations (SNVs) and indels. In the recent past, several research groups reported genetic diagnosis for rare disorders using WES applications while a few used characteristic trio-exome analysis to infer the candidate or driver mutations (*Boycott et al., 2013; Gahl et al., 2012; Jacob et al., 2013*). With human disease and genetic variation studies largely driven by NGS, a paramount challenge would be to explore *de novo* mutations, i.e. those inherited from either father or mother. To check this, parent-child trios/quad WES analysis could be a powerful approach although biases impede identifying potential *de novo* mutations. Nevertheless, a majority of mutations may not transcend from parent to offspring making comprehensive genetic variants to be analyzed for confirming them as causal (*MacArthur et al., 2014*). The trio-based exome sequencing has provided a benefit for identifying *de novo* variants in rare diseases attributing them to be heterozygous/causal mutations (*Zhu et al., 2015*). For rare diseases, although WES analysis often makes assumptions regarding disease inheritance (*de novo* vs. recessive), variant frequency and genetic heterogeneity, it has opened the path towards improved disease management or prognosis and effective therapies. For example, WES trios in schizophrenia patients for recessive genotypes were studied with rare mutations in voltage gated sodium ion channels contributing to the disorder (*Rees et al., 2015*). In another study, Jin *et al*. (2017) have identified the pathogenic mutations with an increased rate of *de novo* mutations in early-onset high myopia (EOHM) patients (*Jin et al., 2017*). Recently, Quinlan-Jones *et al*. have correlated the proband–parent trios to determine the clinical utility of molecular autopsy underlying etiology of structural anomalies (*Quinlan-Jones et al., 2019*).

Congenital Pouch Colon (CPC) is a rare type of high anorectal malformation wherein a part of or entire colon gets dilated in the form of pouch having fistular communication with genitor-urinary tract (*Mathur et al., 2002*). Most of the incidences have been reported from India with cases common to other countries accounted although the males are prone to be largely affected, i.e. male to female ratio of 4:1 (*Gupta and Sharma, 2007*). From WES approaches, we have earlier identified mutations that are causal to CPC and reported candidate missense and nonsense mutations in genes, *viz*. C7orf57, C10orf120, C9orf84 and MUC16, CTC1 respectively (*Mathur et al., 2018*). In a separate study, we have also inferred the role of long non-coding RNAs (lncRNAs) from a WES and identified *lnc*-EPB41-1-1, located in the intergenic regions of EPB41 that is known to be interacting with KIF13A (*Gupta et al., 2018*). In this extended WES study of CPC, we examine the role of *de novo* mutations in a 16 proband/parent trio family comparing the mutations to that of their unaffected parents. Keeping in view of understanding the genetic basis of CPC that could possibly delve into phenotypic variation, an attempt was made to discover a number of *de novo* variants contributing to the heterogeneity. We identified mutations spanning the orphan open reading frames (ORFs), and a few frame shift mutations, besides mutations in GBA3 and HAVCR1 associated with intestine and urogenital tissues.

## Results and Discussion

The downstream analyses leading to the variant calling was done carefully to yield the list of final number of variants. We achieved a mean read depth of 80 in the targeted regions for an average depth of coverage 110x. We generated a norm of 840667 total variants comprising 775262 SNPs and 65405 indels per exome in probands as compared to 736820 SNPs and 63418 indels in unaffected samples from a total of 777216 variants (Figure 1; see methods). Missense variants constituted the large variation followed by loss of function variants in cases when compared to unaffected samples, with an overall 0.35% missense, 0.02% frameshift, 0.004% stop gain and 0.0009% stop lost (Table 1). From the final list of inferring causality, we identified 40 SNPs/ indels with a few non-synonymous mutations as a result of low quality calls (Table 2) which in turn were mapped to 37 single nucleotide variants (SNVs) (Table 3) and 385 copy number variants (CNVs) (Table 4). Given the rare phenotype, the frequency of the confirmed variants was checked for in agreement with that of 1000 genome and ExAC databases. From the reported familial history, the relatedness tests were not felt necessary as the pedigree confirmed correct parenthood for all affected/unaffected samples with the shared homozygosis (Figure 2 and supplementary table 1). Finally, a set of candidate variants inferred from all samples were checked for validation using Sequenom array/plex (Table 5).

**Table 1:** Total variants in cases versus controls

|  | Probands (N=16) | Average variants for Proband | Controls(N=46) | Average variants for Control | Total Cases | % Probands | % Controls |
| --- | --- | --- | --- | --- | --- | --- | --- |
| **Total variants** | 13450685 | 840667 | 3,57,51,971 | 777216.76 | 4,92,02,656 |  |  |
| **SNPs** | 12404198 | 775262 | 3,38,93,726 | 736820.13 | 4,62,97,924 | 25.2104236 | 68.88596827 |
| **Indels** | 1046487 | 65405.4 | 29,04,835 | 63148.587 | 39,51,322 | 2.12689128 | 5.903817469 |
| **Missense** | 172232 | 10764.5 | 4,83,790 | 10517.174 | 6,56,022 | 0.350046144 | 0.983259928 |
| **Frameshift** | 10572 | 660.75 | 30183 | 656.15217 | 40755 | 0.021486645 | 0.061344249 |
| **Stop gain** | 2460 | 153.75 | 6778 | 147.34783 | 9238 | 0.00499973 | 0.013775679 |
| **Stop lost** | 469 | 29.3125 | 1280 | 27.826087 | 1749 | 0.000953201 | 0.002601486 |

**Table 2:**
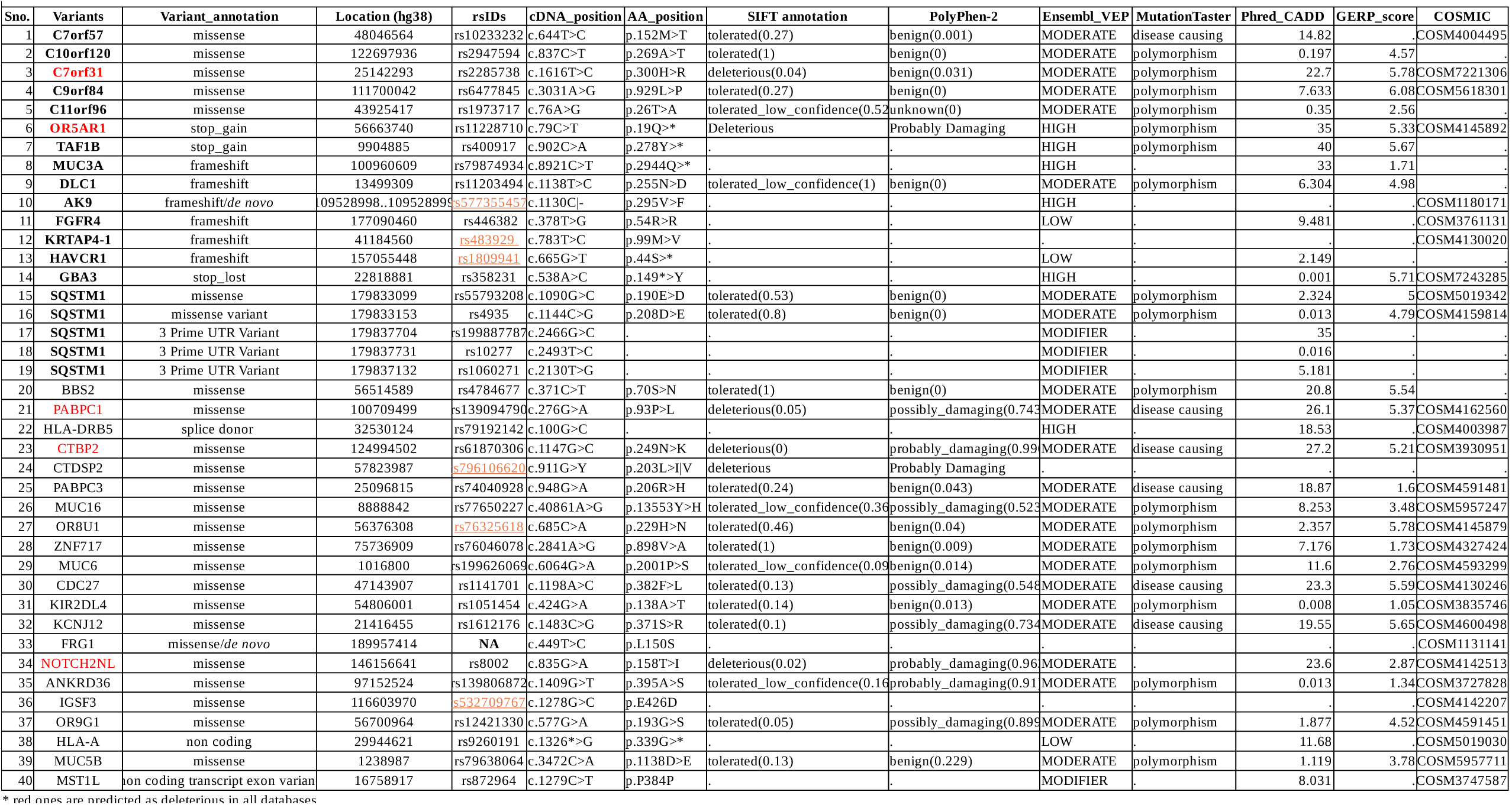
Variants inferred to be causal for CPC from trio-exome analysis

**Table 3:** SNVs identified from trio exome analysis

| Table 3: SNVs identified from trio exome analysis |  |  |  |  |
| --- | --- | --- | --- | --- |
| Sno. | Position | Ref_Allele | Alt_Allele frequency | SNP_id |
| 1 | chr1:16758917 | C | T (2.3170%) | rs872964 |
| 2 | chr1:116603970 | C | G (0.0019%) | rs532709767 |
| 3 | chr1:146156641 | G | T (0.5165%) | rs8002 |
| 4 | chr2:9904885 | C | T (82.6419%) | rs400917 |
| 5 | chr2:97152524 | G | T (0.0148%) | rs139806872 |
| 6 | chr4:22818881 | A | T (82.1699%) | rs358231 |
| 7 | chr5:157055448 | G | T (83.0161%) | rs1809941 |
| 8 | chr5:177090460 | T | G (70.9025%) | rs446382 |
| 9 | chr5:179833099 | G | C (1.7278%) | rs55793208 |
| 10 | chr5:179833153 | C | T (58.5686%) | rs4935 |
| 11 | chr5:179837132 | T | G (0.9383%) | rs1060271 |
| 12 | chr5:179837704 | G | C (0.1700%) | rs199887787 |
| 13 | chr5:179837731 | T | C (59.4508%) | rs10277 |
| 14 | chr6:29944621 | A | G (39.2973%) | rs9260191 |
| 15 | chr6:32530124 | C | T (0.1694%) | rs79192142 |
| 16 | chr7:25142293 | T | C (61.7639%) | rs2285738 |
| 17 | chr7:48046564 | T | C (83.3657%) | rs10233232 |
| 18 | chr7:100960609 | C | T (1.1563%) | rs79874934 |
| 19 | chr8:13499309 | T | C (9.5043%) | rs11203494 |
| 20 | chr8:100709499 | G | A (8.3104%) | rs139094790 |
| 21 | chr9:111700042 | A | G (77.7298%) | rs6477845 |
| 22 | chr10:12269793 | C | T (74.6677%) | rs2947594 |
| 23 | chr10:12499450 | G | C (0.0039%) | rs61870306 |
| 24 | chr11:1016800 | G | A (0.5667%) | rs199626069 |
| 25 | chr11:1238987 | C | A (0.1410%) | rs79638064 |
| 26 | chr11:43925417 | A | G (15.3193%) | rs1973717 |
| 27 | chr11:56376308 | C | T (0.0483%) | rs76325618 |
| 28 | chr11:56663740 | C | T (65.5350%) | rs11228710 |
| 29 | chr11:56700964 | G | T (0.2808%) | rs12421330 |
| 30 | chr12:57823987 | G | C (0.0006%) | rs796106620 |
| 31 | chr13:25096815 | G | A (0.2769%) | rs74040928 |
| 32 | chr16:56514589 | C | T (94.0485%) | rs4784677 |
| 33 | chr17:21416455 | C | T (23.5079%) | rs1612176 |
| 34 | chr17:41184560 | T | C (6.6212%) | rs483929 |
| 35 | chr17:47143907 | A | C (0.0090%) | rs1141701 |
| 36 | chr19:8888842 | A | G (0.1488%) | rs77650227 |
| 37 | chr19:54806001 | G | A (26.8694%) | rs1051454 |

**Table 4:**
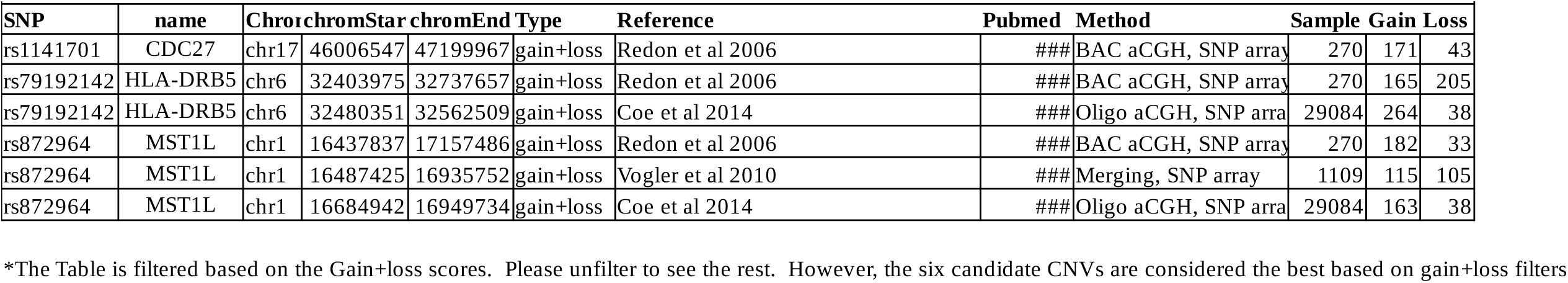
CNVs associated with variants identified from trio-exomes*

**Table 5:** Variant Validation through Sequenom Mass Array plex

| **Rs id** | **nAllele** | **Coverage** | **NoCall** | **Total** | **COMMON** | **HET** | **RARE** |
| --- | --- | --- | --- | --- | --- | --- | --- |
| rs199626069 | 2 | 100% | 0 | 37 | 16 | 5 | 5 |
| rs11228710 | 2 | 100% | 0 | 37 | 16 | 11 | 11 |
| rs77650227 | 2 | 100% | 0 | 37 | 13 | 4 | 4 |
| rs10233232 | 2 | 100% | 0 | 37 | 11 | 9 | 9 |
| rs358231 | 2 | 100% | 0 | 37 | 15 | 7 | 7 |
| rs55793208 | 2 | 100% | 0 | 37 | 9 | 12 | 12 |
| rs2947594 | 2 | 100% | 0 | 37 | 5 | 11 | 11 |
| rs139094790* | 2 | 95% | 2 | 37 | 12 | 4 | 4 |
| rs199887787 | 2 | 100% | 0 | 37 | 12 | 4 | 4 |
| rs2285738 | 2 | 100% | 0 | 37 | 9 | 12 | 12 |
| rs1809941 | 2 | 100% | 0 | 37 | 10 | 12 | 12 |
| rs446382 | 2 | 100% | 0 | 37 | 12 | 9 | 9 |
| rs79638064 | 2 | 100% | 0 | 37 | 7 | 10 | 10 |
| rs139806872 | 2 | 100% | 0 | 37 | 11 | 8 | 8 |
| rs74040928 | 2 | 100% | 0 | 37 | 16 | 5 | 5 |
| rs1060271 | 2 | 100% | 0 | 37 | 14 | 3 | 3 |
| rs577355457 | 2 | 100% | 0 | 37 | 25 | 12 | 12 |
| rs11203494 | 2 | 100% | 0 | 37 | 11 | 7 | 7 |
| rs1973717 | 2 | 100% | 0 | 37 | 12 | 4 | 4 |
| rs532709767 | 2 | 100% | 0 | 37 | 13 | 4 | 4 |
| rs872964 | 2 | 100% | 0 | 37 | 9 | 13 | 13 |
| rs400917 | 2 | 100% | 0 | 37 | 10 | 7 | 7 |
| rs4784677 | 2 | 100% | 0 | 37 | 9 | 10 | 10 |
| rs79874934 | 2 | 100% | 0 | 37 | 8 | 12 | 12 |
| rs79192142 | 2 | 100% | 0 | 37 | 5 | 13 | 13 |
| rs61870306 | 2 | 100% | 0 | 37 | 4 | 16 | 16 |
| rs12421330 | 2 | 100% | 0 | 37 | 13 | 4 | 4 |
| rs1612176 | 2 | 100% | 0 | 37 | 15 | 5 | 5 |
| rs6477845 | 2 | 100% | 0 | 37 | 12 | 12 | 12 |

\*This SNP failed in validation

**Figure 1:**
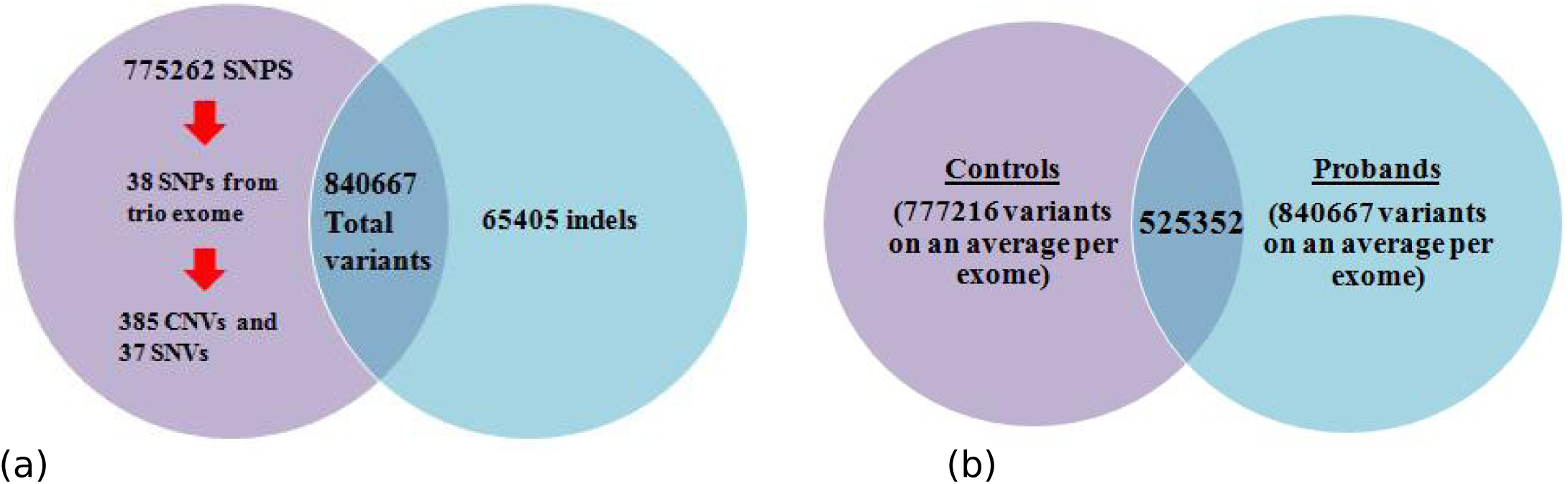
Venn diagram showing variants per exome in (a) probands only and (b) controls versus probands.

**Figure 2:**
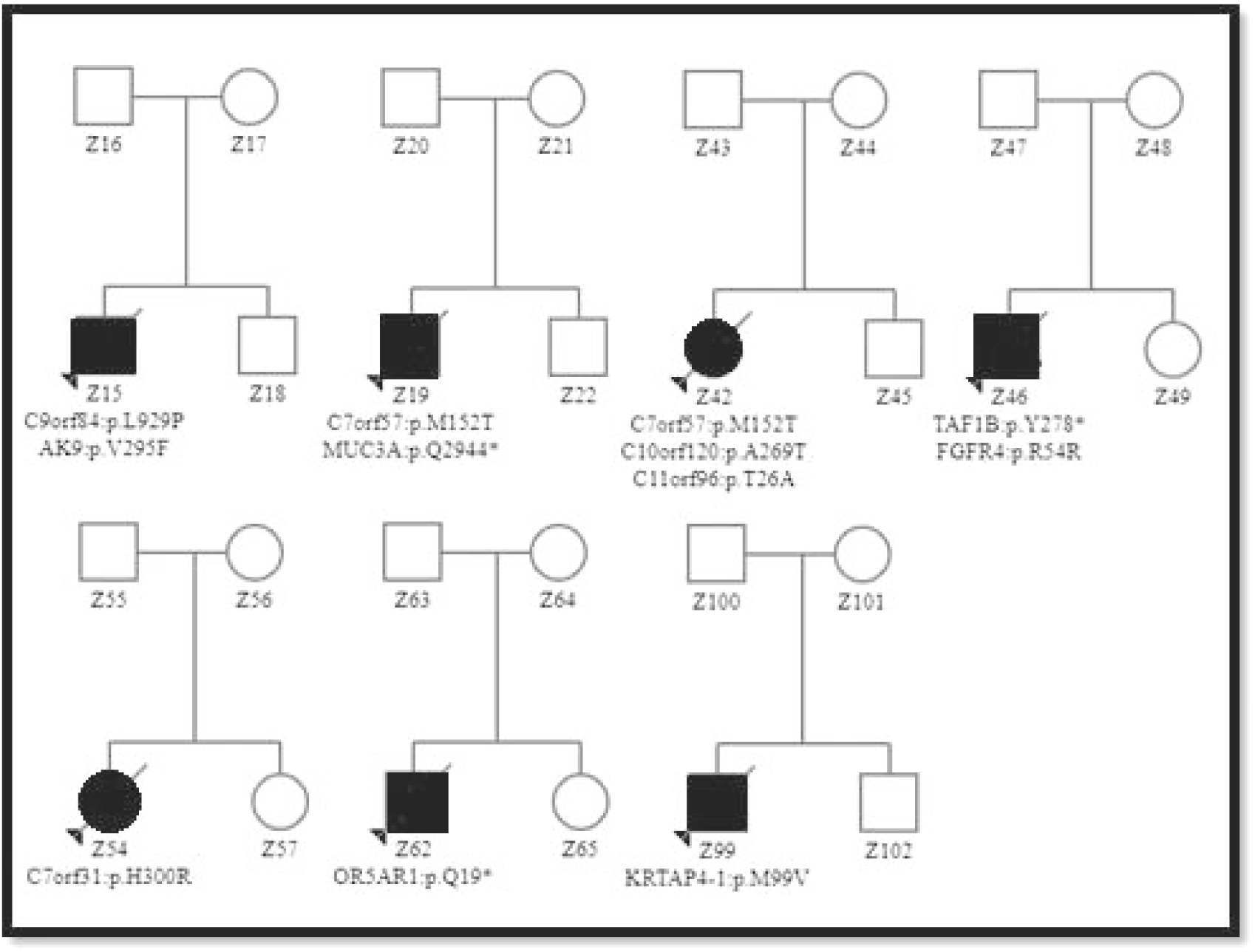
Families with inherited mutations in CPC probands (marked with arrow, please refer supplementary Table 1 for details)

We found two independent lines of evidence from various tools and reached consensus in detecting variants from each samples with all filtering steps. For example, the pathogenic mutations contributing to the each relevant proband were screened first. Altogether, we observed that MUC3A, TAF1B, in addition to the two hypothetical protein-coding genes, *viz*. C10orf120 and C9orf84, contribute to CPC and these could be interesting as they could be novel genes responsible for pathogenesis of CPC. Although from dbSNP, we checked whether or not the SNPs are missense/frameshift variants, we attempted to validate them according to their clinical/pathogenic significance besides checking the heterozygous mutations for minor allele frequencies (MAF<0.01). Among them, 11 variants were already reported in dbSNP (C7orf57, C10orf120, C7orf31, C9orf84, C11orf96, OR5AR1, TAF1B, MUC3A, DLC1, AK9, FGFR4, KRTAP4-1) whereas AK9 was found to be *de novo* in two probands (Z19 and Z99) with another putative *de novo* variant, *viz*. FRG1, which was later found to be a false positive post Sequenom validation. In summary, we observed FRG1 and AK9 as *de novo* variants in CPC-XI and CPC-XII, CPC-XVI families. While FRG1 is a protein coding gene involved in mRNA transport and processing linked with epigenetic regulation of muscle differentiation by regulating histone-lysine N-methyltransferase activity (*Sun et al., 2011*), it is associated with disease pathogenesis and its over-expression leads to development of facio-scapulohumeral muscular dystrophy (FSHD1)-like symptoms. On the other hand, AK9 is involved in nucleotide metabolism pathways and maintains homeostasis of cellular nucleotides (*Amiri et al., 2013*).

One of the important challenges for researchers and clinicians nowadays in investigating rare disorders involves predicting pathogenicity of variants of unknown significance (VUS). With several guidelines mentioned for predicting pathogenicity of variants (*Landrum et al., 2016; MacArthur et al., 2014; Richards et al., 2015*), molecular investigators face a daunting task in considering a rare variant as benign or pathogenic and inferring them to pathogenesis. In rare diseases, where only a minority of population is getting affected and also prevalent to a specific geographical location, identifying and considering VUS will require a thoughtful consideration. In our study, candidate variants such as SQSTM1 (rs10277), GBA3, FGFR4 and HAVCR1 are shown to be reported in urogenital and colon related disorders and are not reported as deleterious in most of the pathogenicity determination databases (Supplementary figure 1). A notable among them, *viz*. sequestome (SQSTM1 or P62) gene encodes a multifunctional scaffolding protein involved in multiple cellular processes (*Sanz et al., 1999*) besides showing mitochondrial integrity, import and dynamics as a discriminating autophagy receptor (*Seibenhener et al., 2013*). In addition, p62/SQSTM1 is expressed ubiquitously in various cell types such as cytoplasm, nucleus and lysosomes (*Pankiv et al., 2010*) and is known to be over expressed in various human genitourinary diseases including colon cancer (*Mohamed et al., 2015*), hepatocellular carcinoma (*Bao et al., 2014*) and prostate cancer (*Kitamura et al., 2006*) (Supplementary Table 2). Although multiple lines of evidence, suggest that apart from VUS, mutations yielding somatic copy number alterations (SCNA) could not be ruled, we found these missense mutations in C7orf57, C9orf84, ORF5AR1, FGFR4, HLA-DRB5, NOTCH2NLA, MUC5B genes associated with CPC. Interestingly, we observed CNVs in CDC27, HLA-DRB5 and MST1L genes with maximum number of gains and losses (Table 4). Particularly, CDC27, a Cdk-inhibitory kinase of the anaphase-promoting complex is known to be associated with colon inactivation.

To confer the mutations associated with pathogenesis, we observed that C7orf31, OR5AR1, PABPC1, CTBP2 and NOTCH2NLA genes harbor deleterious mutations which were screened through SIFT, PolyPhen2, MutationTaster, Ensembl VEP, Phred CADD score (>18) GERP score and COSMIC prediction tools (Table 2). Interestingly, CTBP2 gene which has a heterozygous missense mutation (rs61870306; c.G1147C; p.N249K) was predicted as lethal with Polyphen score of 0.996 showing high conservation among species. Therefore, we considered to model the PDB structure with these mutations showing unfavorable free energy change (ΔΔG) with reduced stability of -1.82 that may affect the protein stability (Figure 3). The interaction between nearby amino acids and other surrounding residues could ultimately reduce the protein stability.

**Figure 3:**
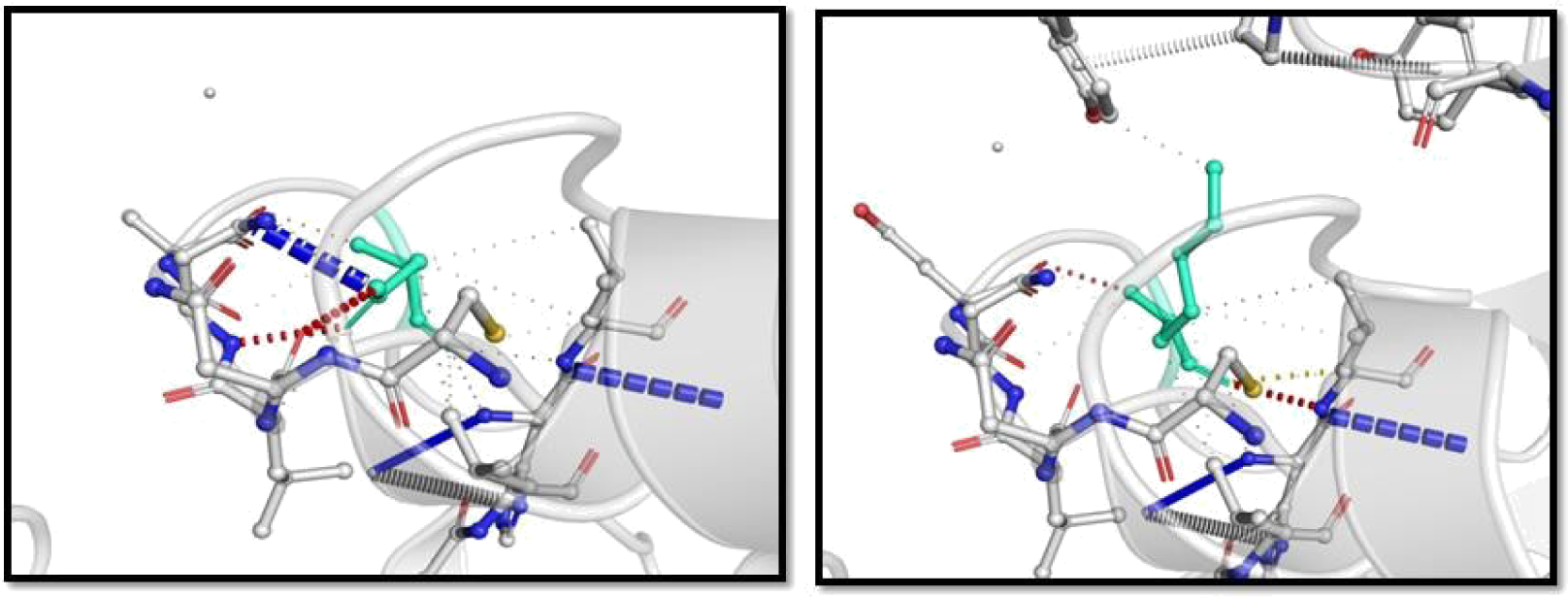
Wild-type and mutant (Asp249Lys) residues are colored in light-green and represented as sticks alongside the surrounding residues.

Dissecting the genetic architecture of a rare disease is certainly an arduous task. Our CPC exome analyses that we have done was intended to fill this gap to detect mutations, SNVs to chromosomal aberrations affecting the focal genes/loci. Assessing these variants in CPC has provided an ample evidence of strata coherent to CPC traits. In our study, at least 4 of the 40 confirmed variants from trio-exome analysis were reported in colon related ailments *viz*. DLC1, HAVCR1, FGFR4 and GBA3. While we have aimed to characterize the variants by employing different approaches, a polygenic model is assumed. This could be compounded with two assumptions (a) capturing exomes and identifying deleterious mutations from high depth of coverage exome and (b) identifying large cohort of mutations that fall in low depth of coverage exomes. Though we observed both classes of genetic variation contributing to the etiology of the disease, inferring proband-parent trios and detecting *de novo* and transmitted genetic variants is quite a challenge. By considering rare variants and adopting a strategy of identifying them in high depth exome, we validated all the 16 trios. Nevertheless, we could not compare the detection yield inherent to these spectrums of patients owing to lack of CPC phenotype and trio-exome studies of similar design. Although previous studies have shown relatively similar methods, they detected medically relevant variants in majority of the diseased phenotypes (*Jin et al., 2017*). Our findings are in agreement with a large number of reports for rare variants, suggesting that cumulative contribution of *de novo* variants across different genes is associated with distinct phenotypes.

One of the interesting findings that have emerged from our study is the role of the known unknowns or hypothetical genes establishing their roles in various diseases. A notable among them is C10orf120 gene which harbors CTCF binding site even as these mutations remain undefined for most disease types including cancer (*Guo et al., 2018*). We observe that there is significant enrichment of indels associated with intestinal/colon related-specific genes as these mutations are widespread in tissues showing chromosomal instability, co-occurring with neighbouring chromosomal aberrations, and are frequent in colon, rectum and gastrointestinal tumours but rare in other diseases. We argue that this mutational disruption associated with CTCF binding sites could be associated with pathogenesis as it appears to be conserved in a majority of CPC probands (Supplementary Table 3). A careful check in Ensembl regulatory feature reveals that the C/T mutation was experimentally validated in mouse (supplementary figure 2) supporting the aforementioned argument. Another orphan ORF, *viz*. C7orf31 also harbours CTCF binding sites and this is in agreement with the fact that the functional pathway analysis showed significant enrichment for biological processes associated with regulation wherein most of our variants are known to be associated with cellular and metabolic pathways (Table 6). Interestingly, we observed that these two ORFs (C10orf120 and C7orf31) are localized to mitochondria and so we asked if there is a plausible role of hypothetical ORFs harboring candidate mutations inherited from mother (rs2947594 and rs2285738). On the other hand, we argue that these mutations are largely autosomal dominant/ *de novo* with a few x-linked or mitochondrial or maternally inherited genes (Supplementary Table 4). Among them, fibroblast growth factor receptor 4 gene (FGFR4) is known to be associated with genitourinary diseases. Although their maternal association cannot be ruled out, we construe that these candidate genes could be promising biomarkers as precursors of CPC. In addition, we screened our variants from Indian Genome Variant Database (IGVDB) and found that they are already reported in Indian subpopulation (*Brahmachari et al., 2005*) and this stratification allowed us to review the patterns influencing common and rare variants. In principle, the rare variants were found to have stronger patterns when compared to the common variants. Thus, there is an inherent need to study the mutations in the known unknown regions which would possibly delve into understanding rare diseases.

**Table 6:**
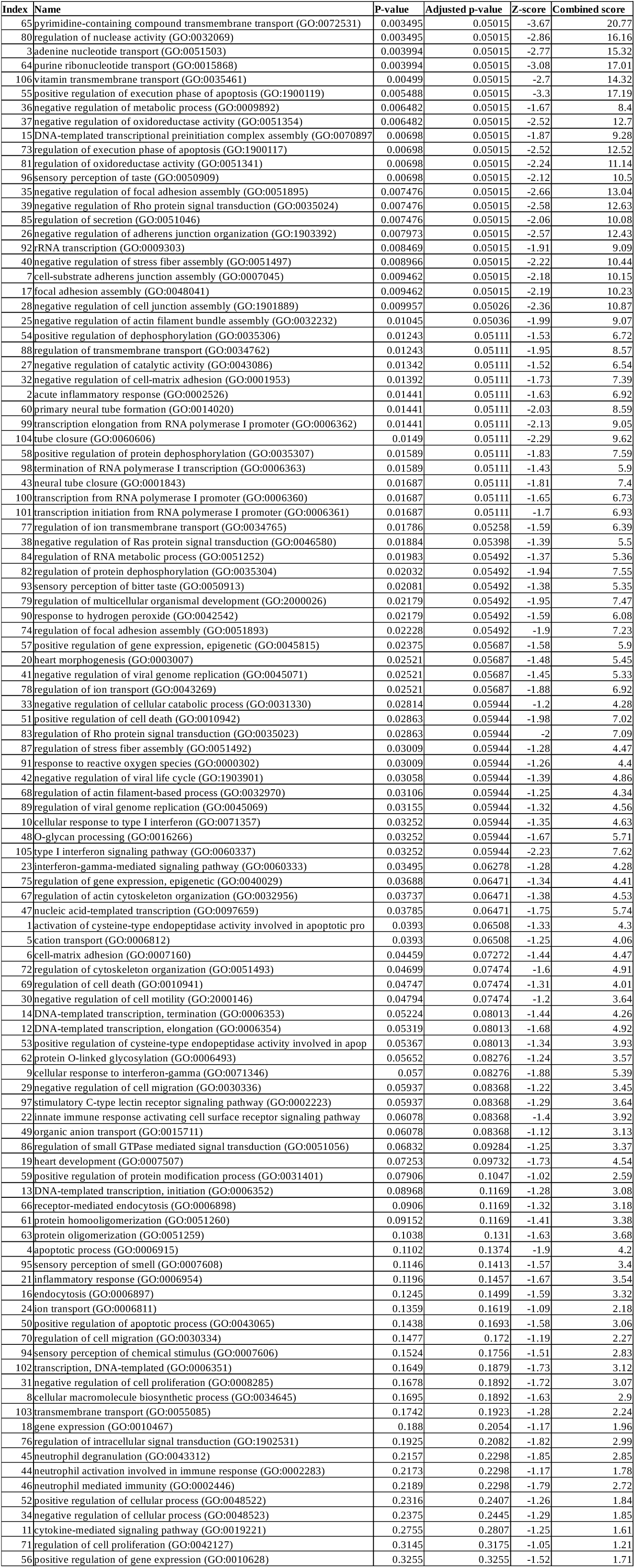
*De novo* gene set enrichment analyses

To gain insights into the role of lncRNAs, we revisited our hypothesis from erstwhile study16 and reconfirmed whether or not NONHSAT002007 was inferred in WES samples with predictions from the NONCODE database. While we did not find mutations in lncRNAs from trio-exome analyses, we argue that the mutations in essential genes tend to be causal for rare diseases paving way for driver mutations with the mutations in non-coding genes suppressed for selective pressure. On a different note, we aimed to evaluate exome enrichment to that of transcriptome and anticipated if any of the variants seen from our WES study could be associated with the differentially expressed genes (DEGs). From the transcriptome pair of CPC type-4 (proband and its unaffected parent), we observed several transcripts, alternative splice variants and fusion genes but none of them could be associated with the causal genes inferred from exome study (Supplementary Table 5). Although we found RGPD2 and RGPD4 genes known to be significantly associated with bowel/colon as among the top enriched, nevertheless it is hypothetical to confirm this inferring global gene expression from just a pair of dataset. This approach, if studied on all samples, we believe, could identify transcripts present at low levels, which in fact could be associated with the pathogenesis of CPC.

In conclusion, we argue that the genetic variability in CPC has had a remarkable significance not only with the parents but also within each proband. The discovery of causal mutations could provide insights into the developmental disorders/anorectal malformation such as CPC and its etiology, which closes the gaps of surgery, taking forward to precision therapy.

## Methods

### Trio selection

The CPC subjects were recruited from the SPJMC (J.K. Lon Hospital), SMS Medical College, Jaipur in accordance with a protocol approved by the institutional ethics committee (IEC) of the hospital. A written informed consent was provided by the parents on behalf of their children. The blood samples were collected from all the probands, parents and unaffected siblings if any. The WES data from a total of 64 samples including 16 affected neonates with their parents and unaffected siblings were used for the study. Although all samples had unaffected sibling samples and data, we restricted our pool of analyses to probands (11 male and 5 female) and unaffected parents/sibling. The methods and pipeline leading to family/quad analyses is summarized in Figure 4 (Supplementary Methods).

**Figure 4:**
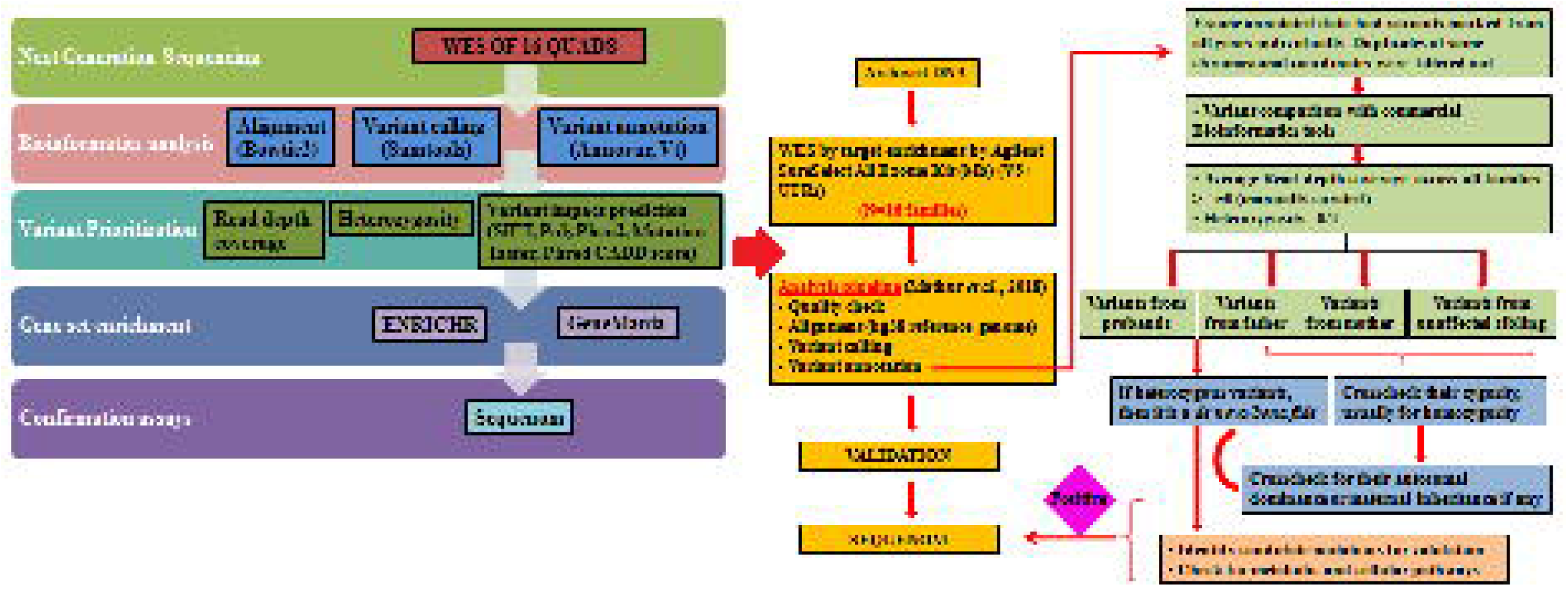
Flow chart for the filtering strategy for trio analysis: A schematic diagram for prediction of causal mutations (Supplementary Methods).

### Variant annotation, filtering and quality control

The details of sequencing and variant calling in CPC subjects have been previously described 15. Briefly, WES was performed on Illumina multiplexed sequencer with paired-end chemistry and 110x effective coverage. Using our in-house developed pipeline, all unmapped sequence reads were aligned to the human reference genome (hg38) and variants were called. The mutations from WES study were manually checked to discover the variants, if any across the samples. To start with, *de novo* variants were interpreted using ‘grep’ and *awk* liner commands, if present in probands but absent in their respective parents (Supplementary information). After manually checking the variants, we confined the prioritization of variants to an average depth of 50 across all the trio samples. Further checking with dbSNP (*Sherry et al., 2001*), ClinVar (*Landrum et al., 2016*) and COSMIC (*Tate et al., 2019*) databases, we used SNP-Nexus (*Chelala et al., 2009*) to filter mutations listed in a cohort of databases, *viz*. SIFT (*Ng and Henikoff, 2003*), PolyPhen-2 (*Adzhubei et al., 2010*), Ensembl Variant Effect Predictor (*McLaren et al., 2016*), MutationTaster (*Schwarz et al., 2014*), CADD (*Kircher et al., 2014*) and GERP (*Cooper et al., 2005*), and confirmed the pathogenic mutations, if they are deleterious in nature. The CNVs and VUS were inferred by mapping the final list of variants to SNP-Nexus. As a final check in reaching consensus for the common variants across all the probands, we compared the variants from multiple bioinformatics tools so as to find *bona fide* variants at the union of intersection of these methods which we construe them to be causal.

### Downstream analyses

To confirm heterozygous mutations, we used *vcftools* package (https://vcftools.github.io) and the mutations in probands were further checked with their relative parents/sibling for heterozygosity transmission. We also looked into the homozygous variants and considered them as causal for CPC where proband is found heterozygous and their respective parents and unaffected sibling if any were homozygous for the specific allele. Enrichment analysis of data was done to calculate inclusive of parameters like binomial probability, hyper geometric distribution (*Huang et al., 2009*). After high throughput screening, we undertook a candidate gene-set analysis based upon significantly enriched fsets or rare mutations specifically in colon related disorders. Seeking novel insights into the disorder, we used pathway analysis based upon Gene Ontology (GO) derived from PANTHER ontology (*Ashburner et al., 2000*) (http://pantherdb.org/tools), EnrichR (*Chen et al., 2013*) (https://amp.pharm.mssm.edu/Enrichr/) annotation terms. Gene Ontology based annotations included Biological Networks Gene Ontology tool (BinGO) (*Maere et al., 2005*), a plug-in for ontology annotation in Cytoscape (*Shannon et al., 2003*) used for ontological analysis in the form of biological, cellular and metabolic processes.

### Identification of transcripts for comparative screening

From RNA-Seq, quality check was ensued after total RNA was isolated and cDNA double strand synthesis on a pair of CPC type-4 samples. The RNA-Seq was performed on Illumina HiSeq 2000 platform with 2x100bp paired end sequencing chemistry which generated ca. 16 million reads. The pair of samples was run through differential gene expression analysis using Cufflinks (*Trapnell et al., 2012*) and DESeq (*Anders et al., 2010*) pipelines and a consensus was reached. For inferring the role of lncRNAs, UVA FASTA software (https://fasta.bioch.virginia.edu/fasta_www2/fasta_list2.shtml#) (v36. 6.8 version) and NONCODE FASTA repository (*Fang et al., 2017*) were downloaded and the intergenic regions of the genes from WES samples were queried. The lncRNA - NONHSAT002007 was identified based on the query coverage e-value < 0.01. The sequences were carefully checked for bidirectional blast hits and the lncRNA was visualized using an Ensembl genome browser for *bona fidelity*.

### SNP Genotyping

For Sequenom genotyping, Multiplexed iPLEX Assay was designed using Assay design suite by Agena Biosciences and MALDI-TOF-MS analysis was performed using Agena Biosciences MassArray Analyzer platform on 29 SNPs in 37 DNA samples (16 probands and 21 controls). This method consists of five steps: PCR amplification, shrimp alkaline phosphatase treatment, single base extension, nanodispensing, and matrix-assisted laser desorption/ionization time of flight (MALDI-TOF) mass spectrometry (*Gabriel et al., 2009*). Data acquisition was automatically performed and mass window of analyte peak observation was set at 4500-9000 Da. Call frequencies, expressed as percentages was calculated for each SNP. For Sequenom, 20 ng of DNA with reference was used to determine 40 SNP calls; however 29 SNPs were validated due to the general failure of SNP calls in, viz. rs483929, rs10277, rs4935, rs8002, rs9260191, rs1051454, rs1141701, rs76046078, rs76325618, and rs796106620 which, therefore, were excluded from our analysis.

## Data Availability

We confirm to the publication ethics and declare no competing interests whatsoever. All WES raw reads are being made available to the federal rare disease portal in India.

## Authors’ contributions

S.G. and P.S. wrote the initial draft and analysed the data. A.K performed Sequenom analysis on discussion with K.M., P.M. and P.S.; K.M. and P.S. conceived the project. P.S. proofread the manuscript before all authors agreed to it.

## Acknowledgements

S.G. acknowledges the support of senior research fellowship from the Council of Scientific and Industrial Research, Government of India (Award #09/892(0002)2015-EMR-I). K.M. and P.S. gratefully acknowledges the support of BTIS-Net, Government of India towards the support of Bioinformatics Infrastructure Facility at BISR, Jaipur.

